# Hospital-acquired infective endocarditis during Covid-19 pandemic

**DOI:** 10.1101/2020.05.17.20101063

**Authors:** Antonio Ramos-Martínez, Ana Fernández-Cruz, Fernando Domínguez, Alberto Forteza, Marta Cobo, Isabel Sánchez-Romero, Ángel Asensio

**Author notes:** Dr. Antonio Ramos. Internal Medicine Department. Hospital Universitario Puerta de Hierro-Majadahonda. C/ Maestro Rodrigo 2. 28222 Majadahonda. Madrid. Spain.

## Abstract

**Background:** The COVID pandemic has had a major impact on healthcare in hospitals, including the diagnosis and treatment of infections. Hospital-acquired infective endocarditis (HAIE) is a severe complication of medical procedures that has shown a progressive increase in recent years.

**Objectives:** to determine whether the incidence of HAIE during the first two months of the epidemic (March-April 2020) was higher than previously observed and to describe the clinical characteristics of these cases. The probability of studied event (HAIE) during the studied period was calculate by Poisson distribution.

**Results:** Four cases of HAIE were diagnosed in our institution during the study period. The incidence of HAIE during the study period was 2/patient-month and 0.25/patient-month during the previous 5 years (p=0.024). Two cases appeared during admission for COVID-19 with pulmonary involvement treated with methylprednisolone and tocilizumab. The other two cases were admitted to the hospital during the epidemic. All cases underwent central venous and urinary catheterization during admission. The etiology of HAIE was *Enterococcus faecalis* (2 cases), *Staphylococcus aureus and Candida albicans* (one case each). A source of infection was identified in three cases (central venous catheter, peripheral venous catheter, sternal wound infection, respectively). One patient was operated on. There were no fatalities during the first 30 days of follow-up.

**Conclusions:** The incidence of HAIE during COVID-19 pandemic in our institution was higher than usual. In order to reduce the risk of this serious infection, optimal catheter care, appropriate use of corticosteroids and interleukin antagonists and early treatment of every local infection should be prioritized during coronavirus outbreaks.

## Introduction

The COVID pandemic has had a major impact on healthcare in hospitals, including the diagnosis and treatment of infections (1–3). In order to reduce the spread of SARS-Cov-2 to patients and hospital staff, priority has been given to the care of life-threatening diseases over non-urgent conditions. In addition, restrictions to the performance of various types of diagnostic procedures and invasive treatments have been recommended (4,5). Experience with prior epidemics has been useful to point out some infection-related issues such as the difficulties to comply both with the use of personal protective equipment (PPE) and the preventive measures for nosocomial infections, as well as the delay in the collection of microbiological cultures and misinterpretation of symptoms and results of diagnostic tests (6–8).

Hospital-acquired infective endocarditis (HAIE) is a severe complication of medical procedures that has shown a progressive increase in recent years (9–11). This progression has been linked to the aging of patients, increased use of venous catheters, parenteral nutrition, hemodialysis, implantation of cardiac devices or cardiac surgery (10–12). To date, no studies have been reported that examine the relationship between the COIVD-19 outbreak and the risk of HAIE.

The aim of the study was to determine whether the incidence of HAIE during the first two months of the epidemic was higher than previously observed and to describe the clinical characteristics of these cases.

## Methods

We performed a prospective study of consecutive adult patients with definite or possible HAIE admitted to our hospital from March 1^st^ to April 30^th^, 2020. IE was defined as per the modified Duke criteria (13). HAIE was defined as either IE manifesting > 48 hours after hospital admission or IE acquired in association with an invasive procedure performed within one month prior to diagnosis (14). Microbiological diagnosis was based on blood or valve culture. Transthoracic and transesophageal echocardiography (TEE) were performed in patients with clinical or microbiological suspicion of IE according to European guidelines, in order to confirm or rule out valve dysfunction and intracardiac complications such as abscesses, vegetations, pseudoaneurysms or fistulae (15). Antibiotic therapy and surgical indications followed the 2015 European Guidelines (15). The probability of studied event (HAIE) during the studied period was calculate by Poisson distribution.

## Results

Four cases of HAIE were diagnosed in our institution during the study period. The mean number of cases of endocarditis per year treated at our center during the last 5 years was 38 ± 3 (mean,SD). Eleven percent of them were considered HAIE. The 4 cases detected during the months of March and April 2020 represent an increase compared to previous 5 years. The incidence of HAIE during the study period was 2/patient-month and 0.25/patient-month during the previous 5 years (p=0.024).

According to Duke’s criteria, the first 3 cases were classified as definite IE whereas the 4^th^ was classified as possible IE (Table 1). Case 1 was diagnosed after the patient has been hospitalized twice in the previous month, for surgical resection of urothelial carcinoma, followed by repair of iatrogenic arteriovenous fistula. Eight days later she was readmitted to a ward specifically dedicated to COVID-19 due to suspicion of having this infection. Five days after admission, IE was diagnosed in the context of moderate heart failure. Case 2 suffered from an aortic prosthetic IE due to *Candida albicans* acquired during admission for aortic valve surgical replacement. During the study period, surgical activity was reduced due to the SARS-CoV-2 pandemic, and the capacity of the surgical ICU was restricted to prioritize COVID-19 cases needing critical care. In any case, the surgical team surgical treatment was rejected because of the high risk of death due to frailty of the patient and the high risk of technical complications during surgery. Cases 3 and 4 presented IE after being diagnosed with COVID and treated with methylprednisolone (more than 1000 mg, each) and two doses of tocilizumab (600 mg, intravenously). Case 3 underwent surgery due to severe mitral insufficiency and evolved favorably. All included patients underwent central venous and urinary catheterization and had received systemic antibiotic treatment during the month prior to the diagnosis of HAIE. Comorbidity, risk factors, infection source, clinical presentation, ETE findings, microbiology and treatment of the patients are shown in Table 1. There were no fatalities during the first 30 days of follow-up.

## Discussion

There is a growing concern about the potentially negative impact of the COVID-19 pandemic on the prevention, diagnosis and treatment of other infectious diseases. (1–3). Our study indicates that the risk of developing HAIE may increase during an outbreak of coronavirus infection. To reduce the risk of HAIE, efforts to improve the prevention and treatment of bacteremia and other hospital-acquired infections potentially related to HAIE should be undertaken during outbreaks of viral respiratory infections.

HAIE cases usually comprise approximately 10% of the IE cases in the literature. The characteristics of the patients included in this small series are similar to those of series published before the COVID-19 pandemic (9–12). Advanced age, prolonged contact with the hospital, the presence of previous heart disease or invasive devices often characterize these patients. With respect to etiology, it should be noted that half of the cases were caused by E. faecalis. This result is in concordance with the increase in enterococcal HAIE frequency that typically affects elderly patients with prior valvular damage or invasive diapositives (28)

Our results represent a remarkable finding because, to our knowledge, no link has been reported between outbreaks of coronavirus and the increase in IE cases so far. In fact, no cases of endocarditis as a co-infection in patients with Covid-19 have been communicated yet. In contrast, co-infection with bacteria and fungi is a proven fact in severe cases of COVID-19 and has been estimated at about 8–10%, with a bacteremia rate of about 6% (17–21). Lymphopenia and immunosuppressive treatments such as corticosteroids and interleukin-6 antagonists such as tocilizumab could favor this complication (18, 22).

Viral pneumonia itself could be considered a risk factor for the development of IE although the occurrence of pneumococcal endocarditis in the context of a respiratory virus pandemic is considered a rare event (23). As has been observed in previous outbreaks, some of our patients may have suffered a delay in the diagnosis of IE due to postponement of blood culture collection, the difficulties in obtaining a TEE for fear of contagion of the viral infection and to the misinterpretation of clinical and microbiological results (7,8). Because of the similarity in clinical presentation between COVID-19 and other infections, appropriate diagnostic tests (including blood cultures) should be promoted when patients show signs suggestive of bacteremia or endocarditis (19). Although TEE is a high-risk procedure for transmission of CoVSARS-2 to health-care workers, because it can generate aerosols, its use should not be restricted only to life-threatening cases, even in the context of the COVID-19 pandemic, as has been proposed (5,13). Providing healthcare workers with the appropriate PPE and facilitating TEE in appropriate cases, a proper diagnosis of IE during outbreaks of coronavirus infection can be done.

With regard to extrinsic risk factors, our patients frequently had invasive devices that increase the risk of developing HAIE such as venous or urinary catheters (14, 17). In fact, the use of PPE could constitute an additional difficulty in the correct maintenance of these devices due to a decrease of the time dedicated to patient care (6). The WHO has insisted on the convenience of verifying sterile insertion in these patients and the need for removal when no longer needed(5). Of note, cases 1, 2, and 4 presented a distinct original source of infection (peripheral catheter infection, sternal wound infection and central venous catheter infection, respectively).

The fact that the study was done in a single center and the small number of cases makes it advisable to await confirmation from larger, multicenter studies before acknowledging the increased risk of HAIE during coronavirus outbreaks. An additional limitation of the present series is that only 2 of the cases were diagnosed with COVID-19 concomitantly with HAIE, complicating further to draw any conclusion; nevertheless, all of them suffered the consequences of the pandemic at the hospital level.

In summary, we have detected an incidence of HAIE higher than usual during the first two months of the COVID-19 pandemic. In order to reduce the risk of this serious infection, optimal catheter care, appropriate use of corticosteroids and interleukin antagonists, early treatment of every local infection and appropriate use of diagnostic techniques in patients with suspected systemic infections should be prioritized during viral respiratory outbreaks.

## Data Availability

All data are available in our data base

## Ethical statement

This study was approval the local Clinical Research Ethics Committee (CEIC). All patients gave their consent to participate in the study.

